# Engaging Community Leaders to Identify Community Assets for Emergency Preparedness

**DOI:** 10.1101/2020.06.16.20129635

**Authors:** Rachael Piltch-Loeb, Dottie Bernard, Beatriz Quinones, Nigel Harriman, Elena Savoia

## Abstract

**Introduction:** Public health and emergency management agencies play a critical role in addressing the needs of vulnerable populations in preparation for and in response to emergencies. Identifying and leveraging community assets is a way to address such needs. Local knowledge about community assets is a valuable and untapped resource that can be leveraged for disaster response and recovery efforts. This study focuses on the development of a process and tools to engage community leaders in sharing their knowledge about their community characteristics and assets useful for emergency planning.

**Methods:** We conducted interviews with community leaders across five study sites with the goal of understanding what type of local knowledge community leaders are able to share in regards to emergency preparedness. Interview questions focused on community challenges and assets that could be useful to plan for emergencies. Based on the interviews’ results we developed and tested a mobile application to generate a directory and map of community assets. Subsequently, we hosted five community meetings and two tabletop exercises to gather feedback, from community leaders and preparedness planners, on the utility of the local knowledge data produced and on the use of the mobile application to gather and share such knowledge.

**Results:** Based on the interviews’ results we identified two main types of local knowledge about community assets for emergency preparedness: communication-based and trust-based local knowledge. Interviews’ data were used to develop a directory of community assets embedded into a mobile application to be used by community leaders to share the location and/or point of access of specific community assets based on their knowledge. Preparedness planners, who tested the application during a tabletop exercise, found it a useful tool to raise awareness about community assets and enhance planning efforts for vulnerable populations.

**Conclusion:** Community leaders’ engagement in preparedness efforts is important to identify community assets that can be leveraged to address the needs of the most vulnerable segments of a community. The use of a directory of community assets, embedded in a mobile application, can enhance information sharing between community leaders and preparedness planners and facilitate the integration of community assets into preparedness efforts.

## Introduction

Public health and emergency management agencies play a critical role in addressing the needs of vulnerable populations in preparation for and response to emergencies. Identifying and leveraging community assets is a way to address such needs. Community assets consist of existing resources vulnerable populations rely upon and engage with at the local level. Public health agencies have used community health assets identification to promote community health in a variety of circumstances. [1, 2] Community health assets include physical, financial, social, and environmental or human resources. [3] These assets can operate as protective and promoting factors to buffer against life’s stresses. [2] We believe that, local knowledge, *the comprehensive system of concepts, beliefs and perceptions generated by community members in a given setting*, is one type of community asset that can be leveraged in preparedness efforts. While practitioners commonly recognize the value of local knowledge in emergency preparedness, [4] there has been limited research on how to define such knowledge and how to collect local knowledge related data. To address this gap, we interviewed community leaders to explore this construct and pilot test the creation of a tool, by the use of mobile technology, to facilitate the collection and mapping of such knowledge in the form of community assets. As such, the objective of this study was twofold: 1) Explore and define what type of local knowledge community leaders can contribute to in preparedness planning, 2) Develop and pilot test a mobile friendly tool to facilitate the integration of community leaders’ local knowledge into preparedness planning.

In this manuscript, we first describe the two conceptual pillars of our study rationale: community-based planning for vulnerable populations and the health asset-based model. Next, we elaborate on the concept of local knowledge and continue by describing study methods and results.

## Background

### Community-based planning (CBP) for vulnerable populations

The Centers for Disease Control and Prevention (CDC) highlights a variety of factors that may influence a person’s vulnerability to an emergency, including socioeconomic status, age, gender, race and ethnicity, English language proficiency, medical conditions and disability.[5] CBP is one strategy employed by disaster management professionals to build local-level capacity and address some of these vulnerabilities. The strategy includes leveraging the knowledge, capabilities and resources of local communities. A successful implementation of CBP requires an understanding of the communities involved and the ways by which these communities function. [6, 7] To date CBP efforts have mainly focused on identifying community needs and vulnerabilities using a deficit-based model where problems in a community are identified solutions are developed relying on resources outside of the community. In our study, we examined the use of a health asset-based model.

### The health asset-based model

The health asset-based model was originally developed for the public health workforce to reorient their thinking on community planning and intervention development. This model is built upon the concept of salutogenesis, a term drawn from medical sociology. Salutogenesis emphasizes factors that support human health and well-being, rather than those causing disease (pathogenesis).[8] The World Health Organization (WHO) defines “health assets” as the resources that individuals and communities have at their disposal that protect against negative health outcomes and/or promote health status.[9] Health assets can operate at the individual, community, or institutional level. In contrast to a deficit-based model, the health asset-based model entails assessing what communities have to offer in building and developing local capacities for reaching their health goals. [2] Community planning and asset mapping are part of the application of a health asset-based model, which focuses on both tangible and intangible community assets. [11] Use of a health assets model can eventually lead to the creation of inventories (or *asset maps*) of the resources and skills available at the community level prior to intervention development. [10] This model has been applied by public health agencies in a variety of contexts such as engaging faith-based leaders to outreach the population, support anti-poverty campaigns, and leverage citizens and institutions in rural communities to reduce barriers to access healthcare services. [10, 12, 13]

### Local knowledge in emergency preparedness

Local knowledge, one type of health asset, is a valuable and untapped resource that can facilitate community problem solving. Local knowledge develops informally over time by individuals and communities, based on experience, and local culture. This may also include the way people observe and measure the environment around them, solve problems, validate new information and establish the processes whereby knowledge is generated, stored, applied and transmitted to others. [14, 15] Many social problems have local origin thus, local knowledge plays a key role in problem identification, definition, legitimization and, most importantly, in finding solutions. The chances of successful policy implementation are low without the understanding and consensus of local actors. [14] Resident involvement in both defining problems and finding solutions is needed to build the legitimacy required to implement policy in an effective manner. [14] In this study, we seek to explore and define the concept of local knowledge in emergency preparedness and tools to facilitate the sharing of such knowledge by community leaders with preparedness planners.

## Methods

We implemented this study in multiple phases. We conducted an initial conceptualization phase by interviewing community leaders that helped us explore the concept of local knowledge in emergency preparedness and types of community assets they could point to as useful for preparedness planning efforts, followed by a tool development phase where we created a directory of community assets in the form of a mobile application. We then conducted a field testing phase where the tool (a mobile application functioning as a directory) was pilot tested. See Figure 1 for a visual representation of the different phases. Below we describe the details of the methods used in each phase.

**Figure 1.**
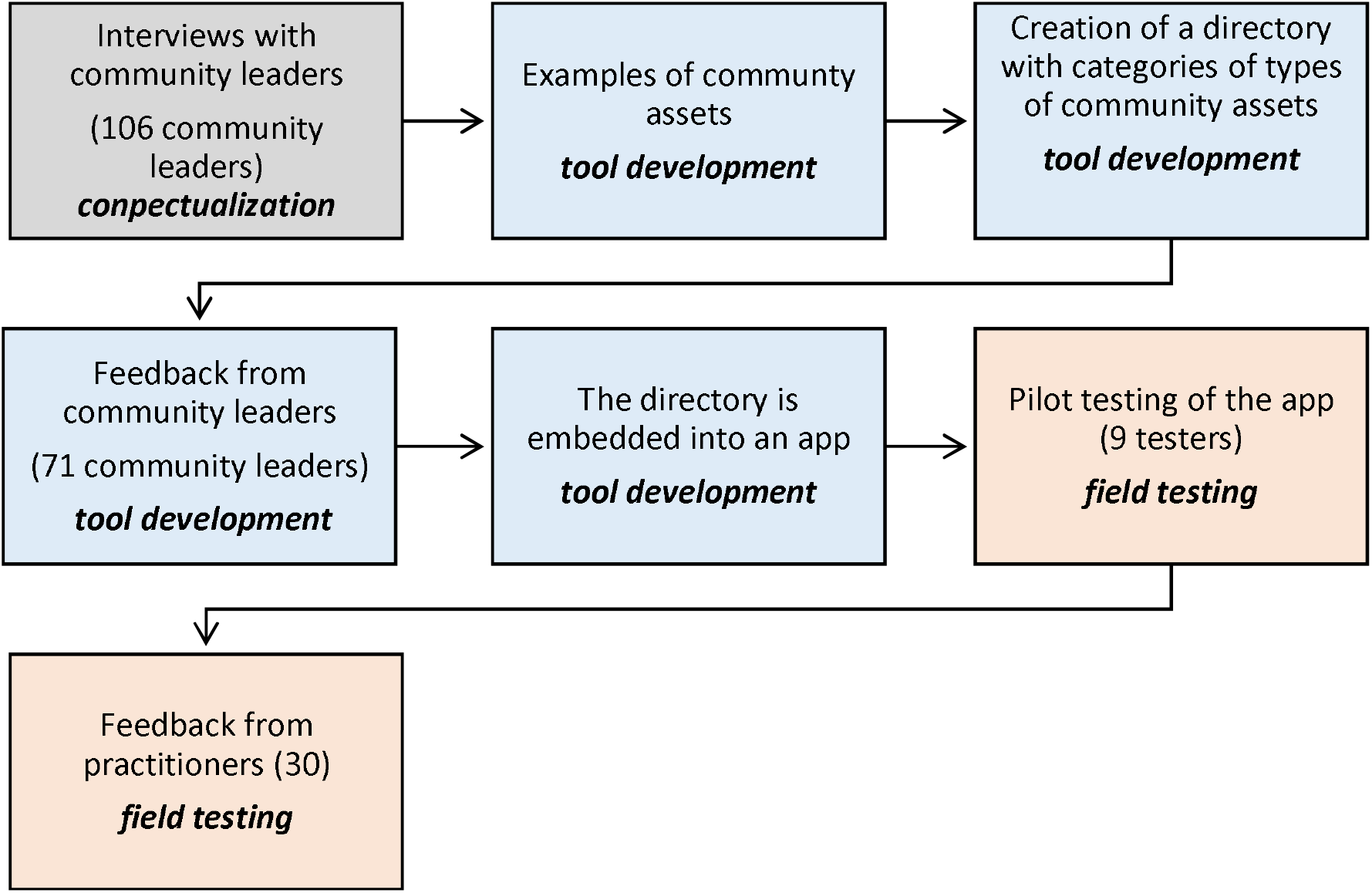
Study implementation steps.

**Figure 2.**
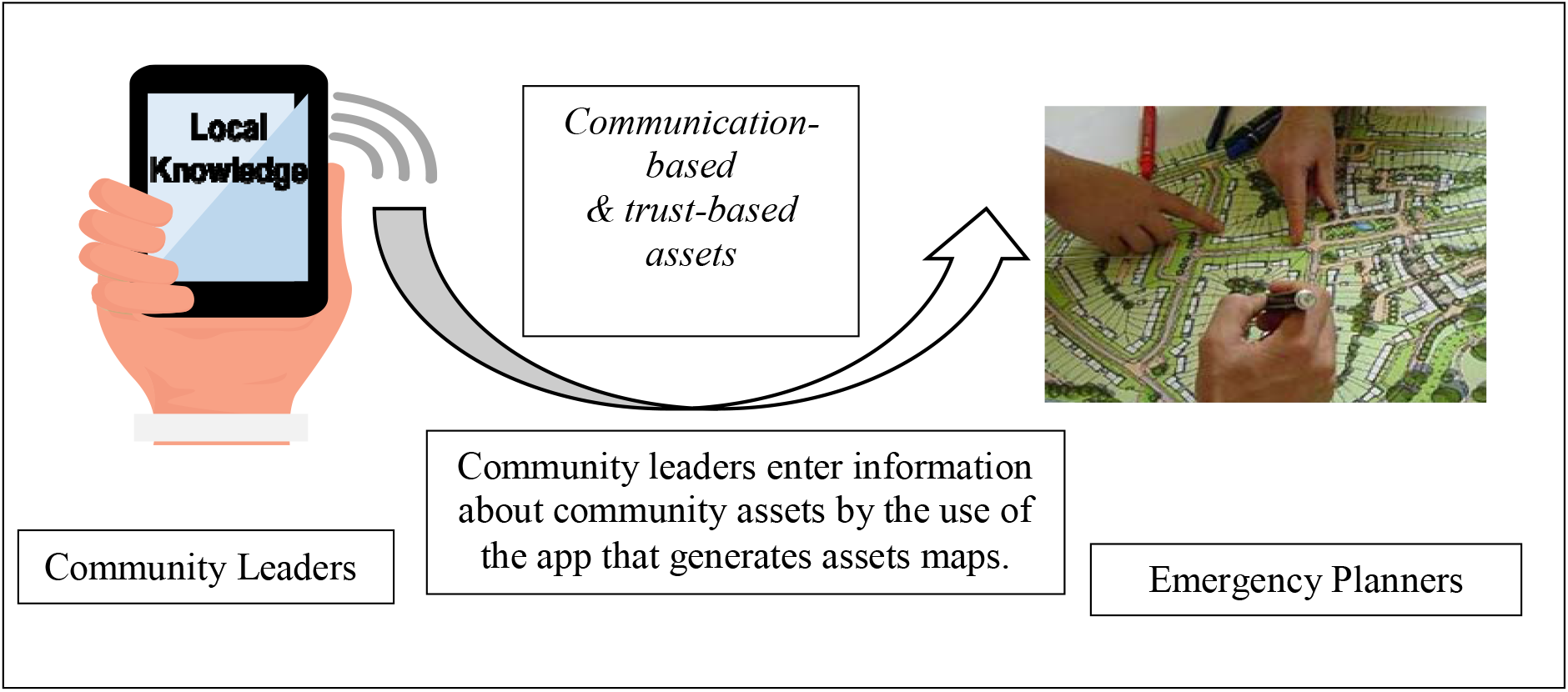
Flow of information between community leaders and emergency planners.

### Conceptualization phase: Interviews with community leaders

With support from pre-identified local partners (community-based organizations) located in the states of Florida, Massachusetts and West Virginia, and the territory of Puerto Rico, we identified and interviewed an initial pool of community leaders. We then applied a snowball technique to recruit additional subjects up to saturation of response content. We recruited community leaders with experience serving vulnerable communities (i.e. living below the federal poverty level, limited English proficiency, ethno-cultural and geographic isolation and drug addiction) and familiarity with emergency preparedness efforts due to either personal experience as a disaster survivor or engagement as a preparedness volunteer (i.e. Medical Reserve Corps volunteers) or as a government official. These leaders could be categorized according to their position and type of interaction with the community in three non-mutually exclusive groups: a) employees of governmental and healthcare organizations, b) members of community-based organizations (i.e. faith-based organizations, volunteer groups, etc.), and c) citizens with substantial civic engagement at the community level. Interviews were conducted in person, following an interview guide and lasted between 45 to 90 minutes. The Harvard Chan School Institutional Review Board deemed the study protocol as exempt.

### Interviewing technique

The interviews were conducted by the use of the convergent interviewing technique. [16] This technique seeks to resolve the dilemma of broad versus specific questions. By the use of this technique data derived from an initial set of interviews are analyzed to inform the development of more specific probe questions in subsequent interviews. Thus, the interview process becomes more and more structured with each subsequent interview. By applying this method, we were able to gather information about specific challenges experienced during emergencies, as well as examples of existing community assets to address such challenges. We started each interview by asking the interviewee to talk about the characteristics of her/his community followed by exploring how community members communicate, where they feel safe and whom they trust during everyday life as well as in emergency situations. Interviews were conducted during the time period August-October 2017 in four languages. The interview guide was first translated from English into the language spoken by the interviewee and then back translated into English for validity purposes. Interviewers had experience in public health practice and were members of the community where the interviews were conducted. We trained interviewers on the use of the interview guide. All interviews were recorded, transcribed and translated into English.

### Interviews’ data analysis

We adopted systematic coding procedures to analyze the interviews’ data using a hybrid method of deductive (pre-set scheme) and inductive (derived from the data) coding. The interview guide included questions on means of communication, trust in organizations engaged in preparedness efforts, personal preparedness, and experience with evacuation and shelter operations. We also assigned codes to reflect both finer distinctions within thematic areas and relationships between topics. Two analysts coded each interview using NVivo v.11 and achieved full agreement on the coding scheme through discussion. A third analyst analyzed the interviews’ transcripts in the language of origin to further validate the results of the coding.

### Tool development and field testing phase

We reviewed the results of the interviews and grouped the examples of community assets named by the interviewees into categories. While the assets cited during the interviews where specific to the selected communities, our goal was to identify and name the categories they belong to in order to develop a directory that could be meaningful to any community. See Table 1. Once the categories and overall structure of the directory was created we hosted community meetings in the same five communities where the interviews were conducted. Seventy-one community leaders, with experience in civic engagement, participated in the meetings and provided feedback on the structure of the directory, and provided suggestions on how to make it more user friendly. Based on such feedback we converted the directory into a mobile application by the use of the Appypie software. [17] A visual representation of the app use is provided in Figure 1.

**Table 1.** Directory categories of community assets.

| Local knowledge area | Directory category<br>(13 codes from the interviews) | Examples derived from the interviews |
| --- | --- | --- |
| **Communication-based assets** | ☐ Community oriented social media channels<br>☐ Ethnic TV stations<br>☐ Ethnic Radio stations<br>☐ Meeting places for large gatherings and places visited by community members on a regular basis<br>☐ Phone trees owned by local organizations<br>☐ Local Translators<br>☐ Organizations & volunteer groups able to conduct door to door outreach | ☐ A Facebook page run by a community member on town news<br>☐ An ethnic TV station streamed online<br>☐ A local radio station hosted in the native language of ethnic community group<br>☐ Worships places, cafe' shops, barber shops, beauty salons<br>☐ A phone tree used by a local church<br>☐ Individuals that volunteer as translators for the community<br>☐ A volunteer group conducting door to door informational campaigns |
| **Trust based assets** | ☐ Organizations with cultural competency and able to maintain confidentiality<br>☐ Organizations with expertise in preparedness | ☐ A volunteer organization or association serving a specific ethnic group<br>☐ A volunteer organization without obligation to report use of illegal drugs<br>☐ A government agency or academic institution with expertise in emergency preparedness |
| **Spatial based assets** | ☐ Spaces culturally accepted and not associated with historical traumatic experiences<br>☐ Spaces that allow for pets<br>☐ Spaces that allow for cultural and religious practices | ☐ A location not associated with historical experience with discrimination or past failures in managing disaster shelters<br>☐ A location that will allow individuals to bring their pets<br>☐ A location that will allow space for religious practice |
| **Personal preparedness assets** | ☐ Organizations that provide assistance to new immigrants | ☐ An educational event hosted by a local organization for new immigrants that will educate them about life in the USA and how to modify food shopping habits when getting ready for emergencies. |

We then engaged nine community leaders to pilot test the app. We instructed the nine testers via webinar on the use of the mobile application, asked to download it on their phone (Androids and IOS) and to use it for a month to enter information about assets in their community across the pre-identified categories and to view the information entered by each-others. Finally, we presented the app and data entered by the community leaders during the pilot test, to a group of preparedness planners in Massachusetts and in Puerto Rico in November 2019 and January 2020. The presentation was embedded into a discussion-based exercise and practitioners were asked to provide feedback on the usefulness of the application for emergency preparedness planning efforts. We presented practitioners with an emergency scenario, consisting of a hurricane in Massachusetts and earthquake in Puerto Rico, and asked them to use the directory of community assets to identify resources that could enhance their public communication capabilities in the preparation phase.

## Results

### Interviews results

We interviewed 106 community leaders belonging to five communities located in three states and one territory: 21 in Florida, 21 in Puerto Rico, 40 in Massachusetts, and 24 in West Virginia. Five were employees of governmental and healthcare organizations, 41 were members of community-based organizations (i.e. faith-based organizations, volunteer groups, etc.), and 60 were citizens with substantial experience in civic engagement at the community level. The coding process led to the creation of 13 major codes, corresponding to categories of community assets that were consolidated into two main themes of local knowledge: communication-based and trust-based local knowledge. See Table 1.

### Local knowledge on communication-based assets

We defined “local knowledge on communication-based assets” as knowledge on assets that relate to preferred means of communication by members of the community. Community leaders talked about how the communities they belong to, which are mostly ethnic communities, (i.e. Somali, Cape Verdean, and Puerto Rican) prefer to receive information through face to face interaction rather than by phone or other means. They reported that knowing about specific places where community members get together (i.e. local churches, restaurants and/or coffee shops) and times of the year during which they are more likely to convene (i.e. religious or country of origin festivities) can be useful for the purpose of hosting preparedness education venues or disseminating information by the use of fliers. Interviewees also highlighted the importance of being aware of age, income and gender differences in the use of specific means of communication. They reported younger members of their community being more inclined to the use of social media, low-income individuals preferring to receive alerts by text message rather than by phone call, and in some ethnic communities (i.e. Somali) women being more likely to receive information from family and friends rather than by attending public events. The interviewees named specific community assets related to the outreach and communication efforts: local translators, mass and social media channels (i.e. ethnic TV and radio stations and community oriented Facebook pages), places where community groups get together or visit on a regular basis (i.e. worship places, building management offices, barber shops and beauty salons), names of organizations that have created text alerts or automated phone call trees, and names of organizations with experience in conducting door-to-door informational campaigns. Community leaders also talked about the need of raising awareness at the community level on the importance of personal preparedness. They reported that availability of seventy-two hour food supplies was unlikely for many community members, mainly due to their low purchasing power which could be supplemented in preparation for an emergency. They also suggested that the best time to get the attention of immigrants on emergency preparedness is when they arrive for the first time in the country and get oriented about life in the USA.

### Local knowledge on trust-based assets

We defined “local knowledge on trust-based assets” as knowledge on assets that relate to organizations, processes and spaces that are trusted by the community. Community leaders talked about the need for the community to be educated about emergency preparedness and that an effective way to do so is by having representatives from government agencies or local academic institutions, with subject matter expertise, host community meetings in close collaboration with organizations and volunteer’s groups trusted by the community. Community leaders pointed to the following criteria as indicative of trusted figures: people/organizations with pre-existing relationships with community groups (defined by ethnicity or specific vulnerability i.e. substance abuse), with cultural competency and with ability to maintain confidentiality regarding immigration status or illicit behaviors (i.e. use of illegal drugs). Community leaders were also asked to describe how citizens experience spaces in the community based on a variety of circumstances. For example, where people do and do not feel physically or emotionally safe (i.e. shelters located in areas affected by previous disasters, places associated with previous experience or symbols of discrimination), the type of transportation they have access to and what basic characteristics “spaces” should have to address their needs. Community leaders talked about how lack of transportation, concerns with safety, privacy in regards to medical conditions and respect of personal and cultural traditions may affect compliance with evacuation orders and sheltering operations. Interviewees also reported several challenges community members could experience if asked to evacuate their homes and go to an emergency shelter such as: having limited access to transportation to reach the shelter site; feel worried to leave their property because of burglary; feel uncomfortable to stay in a location where there are people they do not know; feel incapable to leave their home if the shelter site does not allow for pets; and that they would refuse to go to the shelter if unable to bring what they need to be able to practice their cultural and religious traditions (i.e. a rug to pray). Interviewees then identified community “spaces” that could potentially address some of these challenges. For example they listed facilities, that are not currently named as a formal or municipal disaster shelter, but that could serve for that purpose because recognized by the community as safe and culturally appropriate places; and local organizations with availability of vans that could be used to fulfill transportation needs.

### Tool and field testing results

A total of 71 community leaders participated in the community meetings across the five study sites. Attendees included representatives from public health agencies, youth groups, elderly services, faith-based, healthcare and educational organizations and volunteer groups. They provided general feedback on the categories included in the directory in terms of content and face validity and specific feedback on how the categories were named. Revisions to the tool were made accordingly. Participants solicited and discussed the idea of turning the directory into a mobile application. As a result of these discussions, we created a prototype for the app version of the directory. A demo of the prototype can be found <u>here</u>. A field-testing phase of the app was implemented by engaging nine community leaders. The nine leaders were surveyed after the pilot phase to report on their experience in using the app. They reported that viewing information about the community assets in the app was either easy or very easy, that they did not experience any technical difficulties but that adding information by the use of the crowdsourcing tool was challenging for half of them. Subsequently, we gathered feedback on the usefulness of the directory from 20 practitioners attending the tabletop exercise in Massachusetts and 10 in Puerto Rico, 83% of which had disaster planning responsibilities within their job duties. Overall, 63% of respondents were working in a public health agency, 17% in a healthcare organization and the rest for community-based organizations. All respondents found the data on community assets entered in the directory as valuable information for emergency planning, in particular about communication planning for vulnerable populations. When asked for what types of emergencies they could see themselves using the directory respondents reported pandemic influenza, foodborne illness, severe weather emergencies and any type of emergency planning affecting infrastructures (i.e. water contamination, power outage). In Massachusetts only 3 out of 20 respondents did not see themselves using the directory in any emergency scenario, in Puerto Rico none. When asked to provide feedback on the usefulness of the directory and its mobile version in an open response question, respondents reported that they believed the app technology could be a friendly instrument *“to raise awareness about the need to outreach vulnerable populations, especially among agencies not used to plan for the needs of vulnerable segments of the population*.*”* They also thought the app could be particularly useful for small communities to help them have better access to resources in nearby towns so that community assets could be stored in the same directory and accessible to all users by the use of a common platform. Some liked the fact that the app could function as a portable dataset of information *“much better than flipping through the pages of a plan*”. Most interestingly, practitioners mentioned that this tool could help develop new lines of communication and prompt emergency planners *“take a different look at their community, do things differently and plan in advance to reach vulnerable groups”*. Regarding constructive considerations, participants also pointed out the need for a gatekeeper for the information entered in the directory to *“ensure the information is up to date and accurate”* and that it would be important to be able to download the directory data in anticipation of situations of power outrage.

## Discussion

The literature shows that people affected by disasters may play a crucial role in preparedness and mitigation efforts, but their knowledge is often ignored by the organizations in charge of the response. [20] Prior studies have focused on developing tools to assess how inter-organizational coordination, including coordination with community-based organizations, helps in meeting the needs that individual organizations cannot meet alone. [21] In our work, we sought to understand the types of local knowledge that could be integrated into preparedness efforts and whether an app could facilitate the sharing of knowledge between community leaders, many of which work for community-based organizations and preparedness planners. In this study, we explored and defined local knowledge in preparedness and we provided results from the pilot testing of a mobile application designed to facilitate the integration of such knowledge into preparedness efforts. The mobile application represents a tool by which this information can be systematically shared in a two-way communication approach where community leaders, who are those availing the knowledge and who have better access to vulnerable communities, can coordinate with disaster planners, who are the ultimate users of such knowledge. We recognize the limitations and challenges of our work. The types of knowledge we identified are certainly not exhaustive of all types of local knowledge in emergency preparedness. We identified two areas of local knowledge community leaders could contribute to across five sites having quite diverse community challenges. We then explored the possibility of using an app to systematically gather such knowledge. Yet, in and of itself, a mobile app for emergency preparedness is not novel. A review by Bachmann and colleagues identified 219 apps ranging in purpose and target users. [22] Most apps consist of alerting mechanisms, educational tools, and citizen-to-citizen apps designed for individuals in a given community who are looking to exchange resources during recovery efforts. However, despite the multitude of preparedness apps, there is currently no mobile application focused on information sharing between practitioners and community leaders. Our app is innovative, as it is a crowdsourcing tool for community leaders to share information with the public health practice and emergency management community with the scope of enhancing preparedness planning efforts. We also recognize that there are technical limitations to using an app to capture local knowledge in preparedness planning. Limitations, for example, include security issues related to the data entered into the application. It is important to identify who is entering, vetting and using the data, which leads to three key processes in the use of the tool: the identification of the community leaders that will be engaged in entering the data, the identification of the public health practitioners and emergency managers that will use the data and the identification of a gatekeeper. Having a clear user vetting and protocols guidelines for information entry and access could be a way to enhance security and ensure the functionality of the app. Our data also indicate that, while the app was easy to use to view the data, uploading information is still a task that requires technical skills that not everyone has. Currently, the project team manages and vets the resources and information entered into the app by the community leaders, future iterations will have to consider the right gatekeeper to sustain this process, which may differ according to the community using the application. Finally, the practitioners that provided feedback on the app and type of data that can be collected by the use of this instrument, showed enthusiasm and appreciation for the possibility of including information on local resources they may not be aware of into their planning efforts. In particular, they emphasized how technology can help them change how they think about their community and plan for vulnerable groups. Yet, integrating local knowledge into preparedness efforts requires a political will to do so. When community leaders are included in planning efforts, and local knowledge is valued as much as traditional knowledge, practitioners need to be ready to implement flexible plans that allow for the integration of such knowledge into their decision-making processes.

## Conclusion

Community leaders’ engagement in preparedness efforts is important to identify community assets that can be leveraged to address the needs of the most vulnerable segments of a community. The use of a directory of community assets embedded in a mobile application can facilitate the integration of community leaders’ knowledge of such assets in preparedness efforts and enhance information sharing between community leaders and preparedness planners.

## Data Availability

All data will be made available upon request.

## Acknowledgments

This project is supported by Broad Agency Announcement award number 200-2016-92417 Public Health Emergency Preparedness and Response Applied Research (PHEPRAR) funded by the Centers for Disease Control and Prevention (CDC). The conclusions, findings, and opinions expressed by the authors do not necessarily reflect the official position of the Centers for Disease Control and Prevention, or the authors’ affiliated institutions. We are thankful to many community leaders engaged in this study and organizations supporting us with outreach to the community, in particular to the Somali Development Center in Boston, the Cape Verdean Association in Brockton, MA and to Mr. Alberto Montrond for his work as diplomatic liaison.

